# New genetic loci discovery for Alzheimer’s disease using explainable deep neural networks

**DOI:** 10.1101/2023.12.11.23299839

**Authors:** Christina Gkertsou, Anitha Kugur Parameshwarappa, Abdurrahman Shiyanbola, Zhanna Balkhiyarova, Samaneh Kouchaki, Inga Prokopenko, Vasiliki Lagou, Ayse Demirkan

## Abstract

**Background:** Genome-wide association studies (GWAS) have shed light on various complex diseases and traits, by detecting more than 400,000 associated genetic loci. This number is expected to drastically increase because of the use of novel artificial intelligence methods offering new ways to study the effects of variants. Deep learning using artificial neural networks (ANN) is a sub-field of artificial intelligence, which simulates how the human brain learns. We aimed at assessing the potential of deep learning in human genetic studies of Alzheimer’s Disease (AD) and how these compare to the traditional statistical methods used in GWAS, by simultaneously testing the two approaches on the same dataset, while discovering new genetic loci associated to AD.

**Methods:** To address this aim, phenotypic and genome-wide SNP data from the UK Biobank was analysed on a binary outcome, AD diagnosis, in two different data balance options, of one-to-one and one-to-two case-control datasets, using 2,764 cases vs 2,764 controls and 5,528 controls respectively matched on gender, age, ethnicity, PC1-20 and genotyping array. Genetic data handling and GWAS were performed using PLINK, whereas neural networks were trained using GenNet, a new ANN tool, with the same datasets, separated into training (60%), validation (20%) and test (20%) sets. Neural network layers were determined using biological knowledge, by annotating SNPs to genes and genes to AD related pathways, using ANNOVAR annotations followed by GeneSCF and KEGG.

**Results:** Significant associations were detected between four SNPs linked to two different genes and AD for the 1 to 1 case-control study design and six SNPs linked to four different genes for the 1 to 2 case-control study design by using PLINK. All identified regions have been previously associated to AD. GenNet identified twelve SNPs on seven genes to be associated with AD, all with biological plausibility, achieving an AUC of 0.80 when using three biologically determined layers and 0.73 when using two layers at the neural networks. No common top SNPs were identified between the machine learning and GWAS models.

**Conclusion:** This is one of the first studies attempting to compare the traditional GWAS to more sophisticated state-of-art methods for understanding the genetic architecture of complex phenotypes using the same dataset. More systematic comparisons with such approaches on real data are needed to enable best practises for machine learning in the analysis of genome-wide genetic data.

## Introduction

Alzheimer’s disease (AD) is the most common type of dementia, affecting more than 32 million people worldwide (Breijyeh & Karaman, 2020; Gustavsson *et al*., 2022). It is characterized by progressive neurodegeneration, exhibiting neuritic plaques and neurofibrillary tangles, after accumulation of amyloid-beta peptide (Αβ) in the medial temporal lobe and neocortical structures, and the hyperphosphorylation of microtubule-associated Tau protein in neurons (De-Paula *et al*., 2012; Hampel *et al*., 2021). The incidence of AD seems to become more prominent in people over the age of 65. It is currently the sixth leading cause of death in the United States of America (Alzheimer’s Association, 2022). Its symptoms include memory loss, decline of linguistic and execution abilities, as well as hallucinations, which are often accompanied by neuropsychiatric symptoms, such as depression and apathy (Kumar *et al*., 2022). There have been 152 studies catalogued by the GWAS Catalog and 2588 genetic variants associated with the disease so far (Sollis *et al*., 2023). The genetic variants with the largest effects are the ones linked to the *APOE* region, but other genes, such as *BIN1*, *TNIP1*, and *CLNK*, have also been associated with the disease (Wightman *et al*., 2021). The association of *APOE* to AD is so striking that the E4 allele of the *APOE* locus alone has an area under the curve (AUC) of approximately 0.68 (Escott-Price *et al*., 2018). Even though AD has a big genetic component, environmental factors, such as air pollution and alcohol consumption, are also believed to contribute to the disease (Eid, Mhatre & Richardson, 2019).

Traditional genome-wide association studies (GWAS) have identified associations between more than 400,000 genetic variants and hundreds of human phenotypes, providing insights into the pathophysiology of complex diseases and traits (Andreassen *et al*., 2023; Fabo & Khavari, 2023). Despite these successes, there are still several limitations to be considered (Fabo & Khavari, 2023). One of the most significant ones is the fact that the GWAS approach relies on statistical modelling, testing each single nucleotide polymorphism (SNP) separately, and the identified variants only account for a small proportion of the heritability (Mieth *et al*., 2021). Furthermore, genetic risk scores (GRS) calculated as risk predictors in precision medicine do not account for genetic interactions (Slunecka *et al*., 2021). In addition to that, GWAS analysis usually requires a large sample size to achieve the necessary statistical power (Korte & Farlow, 2013). Due to excessive multiple testing, a stringent threshold of significance is necessary to avoid false positive findings at the cost of losing some important variants with smaller effects. Finally, GWAS findings often lack direct biological interpretation and post-GWAS analyses integrating diverse types of data are necessary for identifying the causal genes and achieving triangulation (Tam *et al*., 2019; Uffelmann *et al*., 2021).

To overcome the aforementioned issues, machine learning algorithms can potentially be used (Doust *et al*., 2022; Mieth *et al*., 2021). Such algorithms can increase performance and power in gene discovery and outcome prediction. Although several machine learning tools have been developed, in this study we focus on a specific type of algorithms that aim to “explain” the decisions made by the machine learning model, hence allowing the easier understanding and interpretation of the results. This is referred to as “explainable artificial intelligence” and when applied on GWAS data, it can not only aid the identification of novel loci, but also provide information on correlation structures and even interactions. Explainable machine learning can unravel which locus and how much it contributes to a phenotype (Mieth *et al*., 2021).

A promising machine learning model is called GenNet developed by van Hilten *et al*. (2021). GenNet is a tool created to tackle the lack of interpretable results and address computational issues related to traditional GWAS. GenNet uses neural networks, as well as prior biological knowledge to create groups of nodes that are connected among layers, reducing the sum of learnable parameters that a fully connected neural network would need. The biological knowledge can be, for example, information on gene annotation, local and global pathways, exon annotation, chromosome annotation, as well as cell and tissue type expression. This way, in the model, the neurons are the biological entities and the weights the effects between the neurons, leading to a biologically interpretable network. An advantage of this method is that it allows the combination of machine learning methods with human biological input (van Hilten *et al*., 2021).

The aim of this study was to use AD data from the UK Biobank and a machine learning model to discover new genetic loci associated with the disease, as well as compare the model’s performance to the traditional GWAS approach (Figure 1).

**Figure 1.**
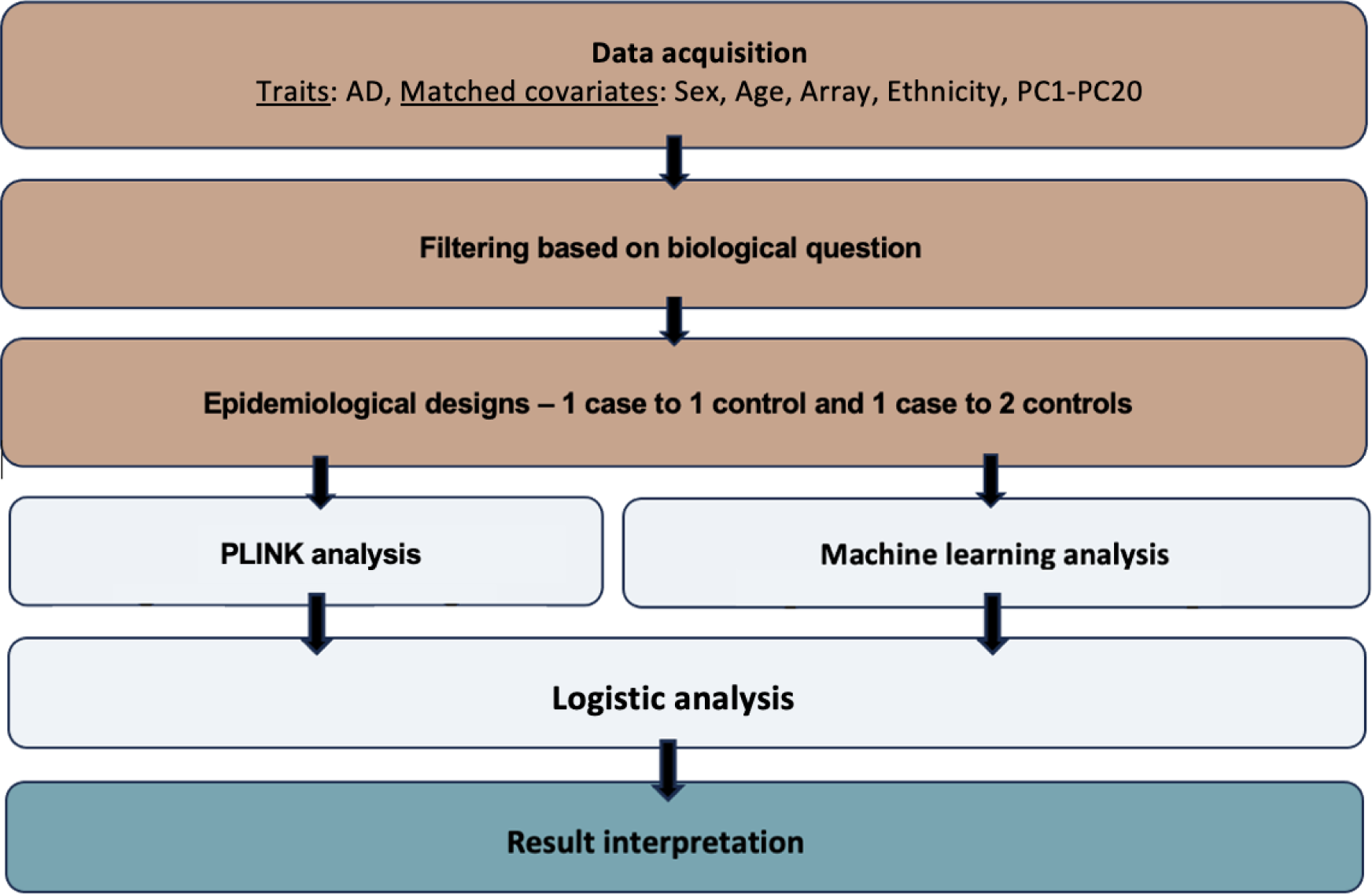
Overview of the analysis. The acquired data were filtered according to the biological question, and afterwards two epidemiological designs - 1 case to 1 control and 1 case to 2 controls - were implemented. The statistical GWAS were performed using PLINK, which follows the conventional statistic-based method of GWAS, and the machine learning analysis was performed using GenNet, which is based on ANN. The last step is the interpretation of the results, which allows the comparison of the two methods.

## Materials and methods

Phenotypes: Data from the UK Biobank was used for the current analysis. The UK Biobank is a large database with medical and genetic data from around 500,000 participants aged from 40 to 69 when recruited, from the United Kingdom (Bycroft *et al*., 2018). This database facilitates research on the prevention, diagnosis, and treatment of a variety of diseases (UK Biobank, 2023). Initial data collection and publications were done during the years 2006 to 2010 with more data and information being recorded since then as follow-ups for different health-related outcomes (Sudlow *et al*., 2015).

The UK Biobank medical dataset used for the current study was the hospital data for Alzheimer’s disease (ID: 131036, ICD10: G30*). The reported number of UK Biobank participants was 4,473 for AD. Age (ID: 21022), sex (ID: 31), genotyping array (ID: 263), ethnicity (ID: 21000), and the first 20 principal components (ID: 22009.0.1 – 2.0) were also extracted to be used as covariates to match on cases and controls in the analysis. The data was published after written consent of the participants and in accordance with the UK Biobank Ethics Advisory Committee. The participants were all registered with the UK National Health Service (NHS) (Sudlow *et al*., 2015).

Matching between AD cases and controls was conducted using the MatchIt package in R (Greifer, 2023). We matched on sex, age, array, ethnicity, and PC1-PC20. A 1 case to 1 control and a 1 case to 2 controls study design ratios were implemented. The final dataset of the 1 case to 1 control study design included 5,528 individuals and the 1 case to 2 controls study design included 8,292 individuals (Table 1).

**Table 1.** Summary of the UK Biobank phenotypic data of interest, grouped by gender.

| 1 case : 1 control design |  |  |  |  |  |  |
| --- | --- | --- | --- | --- | --- | --- |
|  | Males |  |  | Females |  |  |
|  | All | Cases | Control | All | Cases | Control |
| Age years<br>(mean [s.d.]) | 65.12 (4.3) | 65.15 (4.1) | 65.09 (4.4) | 65.26 (4.2) | 65.23 (4.4) | 65.29 (4) |
| AD | 2862 | 1425 (49.8%) | 1437 (50.2%) | 2666 | 1339 (50.2%) | 1327 (49.8%) |
| Ethnicity | Caucasian | Caucasian | Caucasian | Caucasian | Caucasian | Caucasian |
| Array | <u>UK Biobank</u><br>Axiom: 2555<br>(89.3%)<br><u>UK BiLEVE</u><br>Axiom array:<br>307 (10.7%) | <u>UK Biobank</u><br>Axiom:1275<br>(89.5%)<br><u>UK BiLEVE</u><br>Axiom array:<br>150 (10.5%) | <u>UK Biobank</u><br>Axiom: 1280<br>(89.1%)<br><u>UK BiLEVE</u><br>Axiom array:<br>157 (10.9%) | <u>UK Biobank</u><br>Axiom: 2364<br>(88.7%)<br><u>UK BiLEVE</u><br>Axiom array:<br>302 (11.3%) | <u>UK Biobank</u><br>Axiom: 1177<br>(87.9%)<br><u>UK BiLEVE</u><br>Axiom array:<br>162 (12.1%) | <u>UK Biobank</u><br>Axiom: 1187<br>(89.4%)<br><u>UK BiLEVE</u><br>Axiom array:<br>140 (10.6%) |
| N | 2862 (51.8%) |  |  | 2666 (48.2%) |  |  |
| 1 case : 2 controls design |  |  |  |  |  |  |
|  | Males |  |  | Females |  |  |
|  | All | Cases | Control | All | Cases | Control |
| Age years<br>(mean [s.d.]) | 65.04 (4.3) | 65.15 (4.1) | 64.99 (4.4) | 65.35 (4.1) | 65.23 (4.4) | 65.41 (4) |
| AD | 4286 | 1425 (33.2%) | 2861 (66.8%) | 4006 | 1339 (33.4%) | 2667 (66.6%) |
| Ethnicity | Caucasian | Caucasian | Caucasian | Caucasian | Caucasian | Caucasian |
| Array | <u>UK Biobank</u><br>Axiom: 3832<br>(89.4%)<br><u>UK BiLEVE</u><br>Axiom array:<br>454 (10.6%) | <u>UK Biobank</u><br>Axiom: 1275<br>(89.4%)<br><u>UK BiLEVE</u><br>Axiom array:<br>150 (10.6%) | <u>UK Biobank</u><br>Axiom: 2557<br>(89.4%)<br><u>UK BiLEVE</u><br>Axiom array:<br>304 (10.6%) | <u>UK Biobank</u><br>Axiom: 1177<br>(71.7%)<br><u>UK BiLEVE</u><br>Axiom array:<br>464 (28.3%) | <u>UK Biobank</u><br>Axiom: 1177<br>(87.9%)<br><u>UK BiLEVE</u><br>Axiom array:<br>162 (12.1%) | <u>UK Biobank</u><br>Axiom: 2365<br>(88.7%)<br><u>UK BiLEVE</u><br>Axiom array:<br>302 (11.3%) |
| N | 4286 (51.7%) |  |  | 4006 (48.3%) |  |  |

Genotypes: Two arrays were used for the genotyping of the individuals. 438,195 participants were genotyped with the UK Biobank Axiom Array and 49,932 were genotyped with the UK BiLEVE Axiom array. Around 850,000 variants were typed directly (Bycroft *et al*., 2018; UK Biobank, 2023). The genotypic data underwent quality control, phasing, and imputation of approximately 96M genotypes. The latter was achieved by using computationally efficient methods, alongside the use of the HRC and UK10K reference panels. Information on the quality control, phasing, and imputation of the data is available in the paper of Bycroft *et al*. (2018) titled “The UK Biobank resource with deep phenotyping and genomic data” (UK Biobank, 2023; Bycroft *et al*., 2018).

The imputed genotypic data collected from the UK Biobank initially consisted of 96 million SNPs from 22 chromosomes from half a million individuals, summing up to 2.5 terabytes. SNPs with minor allele frequency <0.05 were excluded as for these SNPs imputation might not be accurate. Pruning of data was done in PLINK using the --indep-pairwise 50 5 0.05 command. This way, only independent genetic variants were kept, leading to data redundancy and reduction of the computational expense for ANN analyses.

An additive model was implemented in PLINK 1.07 to test for association (“-assoc”) with AD (binary trait) (“-logistic”). SNPs that achieved a *p*-value lower than 5 × 10^−8^ were considered as statistically significant (Purcell, 2010). After running the association analysis for each autosomal chromosome separately, files were merged and QQ and Manhattan plots were generated. As the lambda (λ) factor showed slight inflation of 1.02 for the 1 case to 1 control design and 1.02 for the 1 case to 2 controls design the chi2 and corresponding p-values were corrected by these genomic inflation factors.

To run GenNet, three files were required: a genotype.h5 file, a subject.csv file and a topology file, as instructed by van Hilten *et al*. (2021). The genotype file is a genotype matrix, with each row showing participant ID. Each column is a feature, for instance a genetic variant. These files were generated by direct conversion of the genotype fam files from PLINK, using the –convert function built in which GenNet which operates in Python (3.9). The subjects file consisted of a minimum of three columns. The first one was named as “patient_id” and included the participant IDs. The second column was named as “labels” and indicated the phenotype. The third column was named as “set”, indicating which subjects would be used for training, validation, test, and which ones should be ignored (1 = training set, 2 = validation set, 3 = test, 4 = ignored). The split of the training, validation and test set was 60/20/20 respectively. Finally, the topology file is the description of the network. The rows indicate the path from the input to the output node, hence the connections of the network (van Hilten *et al*., 2021). For such connections to be built we first connected all SNPs in the plink .bim file to genes (layer 1) using two approaches. First, we annotated each SNP with its most implicated gene reported by Open Target Genetics and identified by integrating genetic and functional genomics features. Second, for variants not annotated by this approach, the nearest gene given by ANNOVAR annotation was hypothesized to be the most implicated one (Ghoussaini et al., 2021; Wang, Li & Hakonarson, 2010). A mapping between genes and pathways (layer 2) was made using GeneSCF and the KEGG databases (Subhash & Kanduri, 2016; Kanehisa & Goto, 2000).

To identify the genes that are potentially linked to the SNPs identified from the two different approaches, Open Targets Genetics was used for the SNPs that had an rsID (Mountjoy *et al*., 2021; Open Targets Genetics, 2023). The Genome Data Viewer by the National Library of Medicine was used to detect the closest genes to the SNPs that had no assigned rsID or had an rsID not found on Open Targets Genetics (Rangwala *et al*., 2020; NCBI, 2023; Mountjoy *et al*., 2021). For such SNPs, it was hypothesized that the closest genes by distance were implicated by the detected variant (Brodie, Azaria & Ofran, 2016). Open Targets Genetics use a variant-to-gene pipeline that integrates evidence from different data types and sources, such as the literature, the UK Biobank, quantitative trait loci experiments, in silico functional prediction, chromatin interaction experiments and distance. Statistical fine-mapping is used to extract an evidence score and detect the association signals and the genes each variant is linked to (Mountjoy *et al*., 2021).

## Results

PLINK analyses showed 4 SNPs being significantly associated with AD (p < 5 × 10^−8^) for the 1 control to 1 case epidemiological design and 6 SNPs for the 1 case to 2 controls epidemiological design. Their p-values were corrected to account for inflation. Interestingly, all SNPs that reached this threshold are located on chromosome 19, which contains loci, such as apolipoprotein E (*APOE*) on its long arm. *APOE* is known to be the strongest risk factor for AD (Moreno-Grau *et al*., 2018).

At the 1 control to 1 case study design, four SNPs were found to be significantly associated with AD (Table 2). From these, two of them, rs10410835 and rs11666329, are functionally linked to nectin cell adhesion molecule 2 (*NECTIN2*), a gene found in the *APOE* region (Kulminski *et al*., 2018). *NECTIN2* is expressed in regions of the brain, having multiple functions, such as contribution to homeostasis of astrocytes and neurons, as well as the formation of synapses. Other SNPs in this genetic region have been previously shown to be associated with AD (Mizutani *et al*., 2022). The rs10629382 variant is associated with B-cell CLL/lymphoma 3 (*BCL3*), a proto-oncogene candidate that has been associated with late-onset familial AD and other cognitive impairment-related diseases (Nho *et al*., 2017). It is upregulated by 27% in the brains of AD patients (Li *et al*., 2018). The 19:45328407_GAC_G variant, based on its distance from the nearest genes, is probably linked either to *NECTIN2* or the basal cell adhesion molecule (Lutheran blood group) (*BCAM)* gene. Both genes are found in the *APOE* locus and are associated with AD (Kulminski *et al*., 2018).

**Table 2.** Top 10 SNPs detected by PLINK association analysis in the AD dataset, arranged by chromosome and position. Only four SNPs (bold) are significantly associated with AD, based on the threshold of p < 5 × 10^−8^. P-values corrected by genomic control (GC) to account for inflation. CHR: chromosome, Position: position of the SNP based on GRCh37, A1: tested allele (minor allele by default), OR: odds ratio, STAT: coefficient t-statistic. Genes refer to the genes that are most likely to be implicated by the SNPs based on Open Targets Genetics and NCBI.

| CHR | SNPs | Position | A1 | OR | STAT | P-value | Genes |
| --- | --- | --- | --- | --- | --- | --- | --- |
| 14 | rs55942844 | 43856713 | C | 1.1820 | 4.401 | 1.28e-05 | *HNRNPUP1* |
| 15 | rs11635698 | 63478573 | A | 1.1930 | 4.590 | 5.35e-06 | *RAB8B* |
| 15 | rs1017546 | 63566702 | C | 1.1840 | 4.411 | 1.22e-05 | *RAB8B* |
| 15 | rs1043256 | 63616479 | G | 1.1860 | 4.487 | 8.64e-06 | *RAB8B* |
| 17 | rs7221678 | 61579612 | T | 1.1960 | 4.712 | 2.99e-06 | *ACE* |
| 19 | **rs10629382** | **45253542** | **CTTTG** | **0.7963** | **-5.635** | **2.32e-08** | ***BCL3*** |
| 19 | **19:45328407_GAC_G** | **45328407** | **G** | **1.2430** | **5.572** | **3.31e-08** | ***BCAM*,**<br>***NECTIN2*** |
| 19 | **rs11666329** | **45354296** | **G** | **0.7853** | **-6.142** | **1.14e-09** | ***NECTIN2*** |
| 19 | **rs10410835** | **45358353** | **C** | **1.3980** | **8.439** | **3.21e-17** | ***NECTIN2*** |
| 21 | rs2829970 | 27257548 | A | 0.8415 | -4.456 | 9.99e-06 | *APP* |

From the study design with the 1 case to 2 controls ratio, 6 SNPs were identified (Table 3). Three of them 19:45328407_GAC_G, rs10629382 and rs10410835 were also found in the 1 case to 1 control study design. The top variant was rs405509, which is known to be located at the *APOE* promoter and is a widely reported risk factor for AD (Ma *et al*., 2016). It is linked to cognitive impairment of the elderly and can affect brain structure as well (Ye *et al*., 2023; Ma *et al*., 2016). Another variant linked to *NECTIN2* was rs57537848. The rs8106813 variant has been previously associated to AD and is linked to the apolipoprotein C1 pseudogene 1 (*APOC1P1)* (Marioni *et al*., 2018). The *APOE* haplotype region is physically interacting with *APOC1P1* (Zhou *et al*., 2019). The latter is associated with the development of late-onset AD (Shigemizu *et al*., 2023).

**Table 3.**
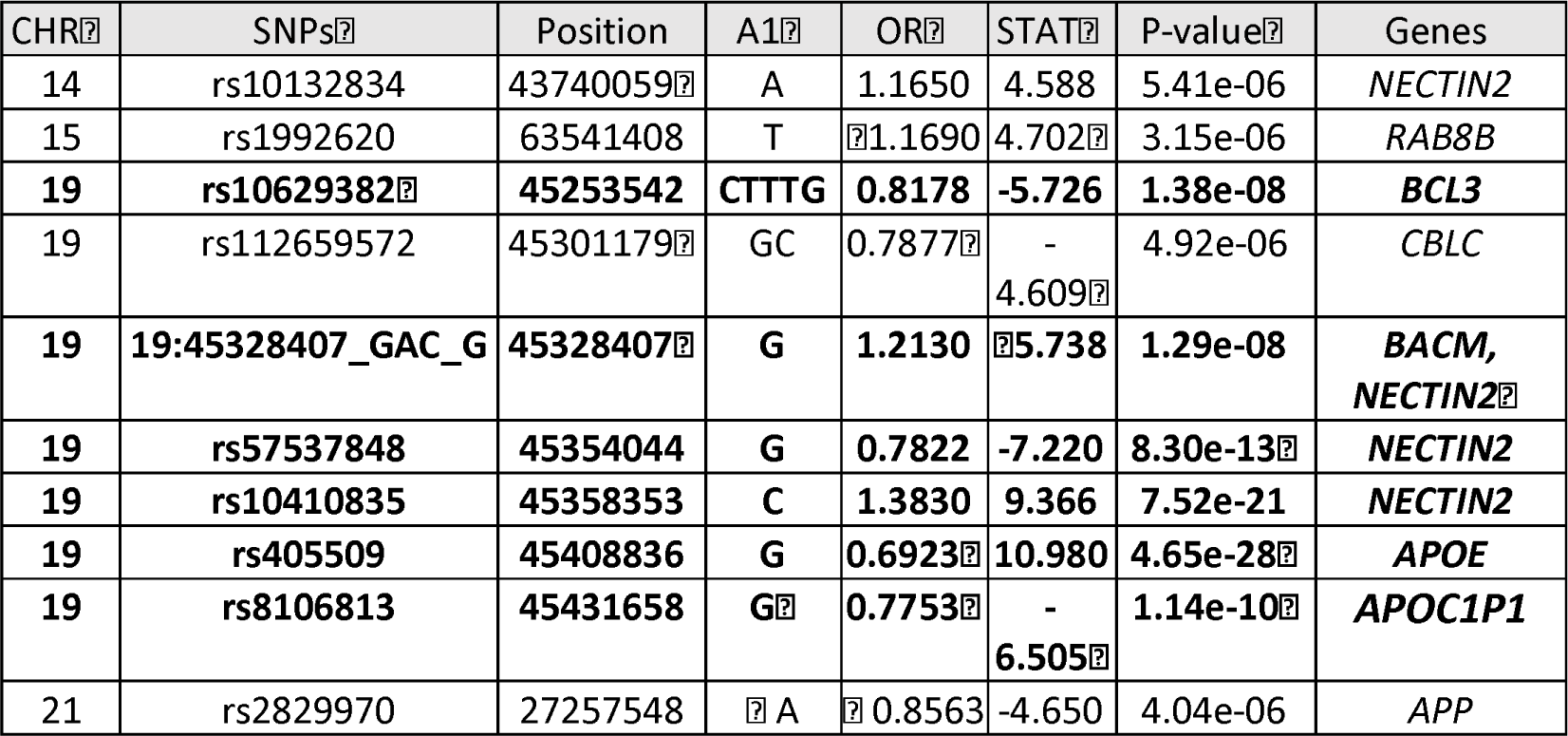
Top 10 SNPs detected by PLINK association analysis in the AD dataset, arranged by chromosome and position. Only six SNPs (bold) are significantly associated with AD, based on the threshold of p < 5 × 10^−8^. P-values corrected by genomic control (GC) to account for inflation. CHR: chromosome, Position: position of the SNP based on GRCh37, A1: tested allele (minor allele by default), OR: odds ratio, STAT: coefficient t-statistic. Genes refer to the genes that are most likely to be implicated by the SNPs based on Open Targets Genetics and NCBI.

To test the associations using GenNet we initially used a 1 case to 4 controls study design. A three-layered model with SNPs, gene annotation and pathway annotation and a two-layered model only with SNPs and gene annotation were implemented.

The three-layered model performed very well for the binary trait. The predictive performance for AD was very good, despite its highly polygenic nature, with an area under the curve (AUC) of 0.80 for the validation set and 0.79 for the test set. The specificity of the test was 0.99, and the sensitivity was 0.028, showing that the model predicted well the negative samples, but not the positive results.

The SNPs with the highest importance values were rs6599892 at chromosome 15 and rs10784050 at chromosome 12 (Figure 2). Although the genes and pathways for those were not identified by the model, the rs10784050 variant is functionally linked to ALG10 Alpha-1,2-glucosyltransferase B (*ALG10B)* (Open Target Genetics, 2023). *ALG10B* encodes a microglial Αβ Response protein. Molecular alterations in microglia, especially because of significant loss of microglial phagocytic function, is a very common characteristic of AD. *ALG10B* might be indicating microglial changes in *early stages of* Αβ *deposition* (Monasor *et al*., 2020). The rs6599892 variant is located on the rhophilin Rho GTPase binding protein 2 pseudogene 1 (*RHPN2P1)*. *RHPN2P1* is expressed in the hippocampus and given the fact that RhoA GTPases play a role in neuron and glial cell signaling, it is hypothesized that it might also have a role in AD (Parcerisas *et al*., 2014; Schmidt *et al*., 2022).

**Figure 2.**
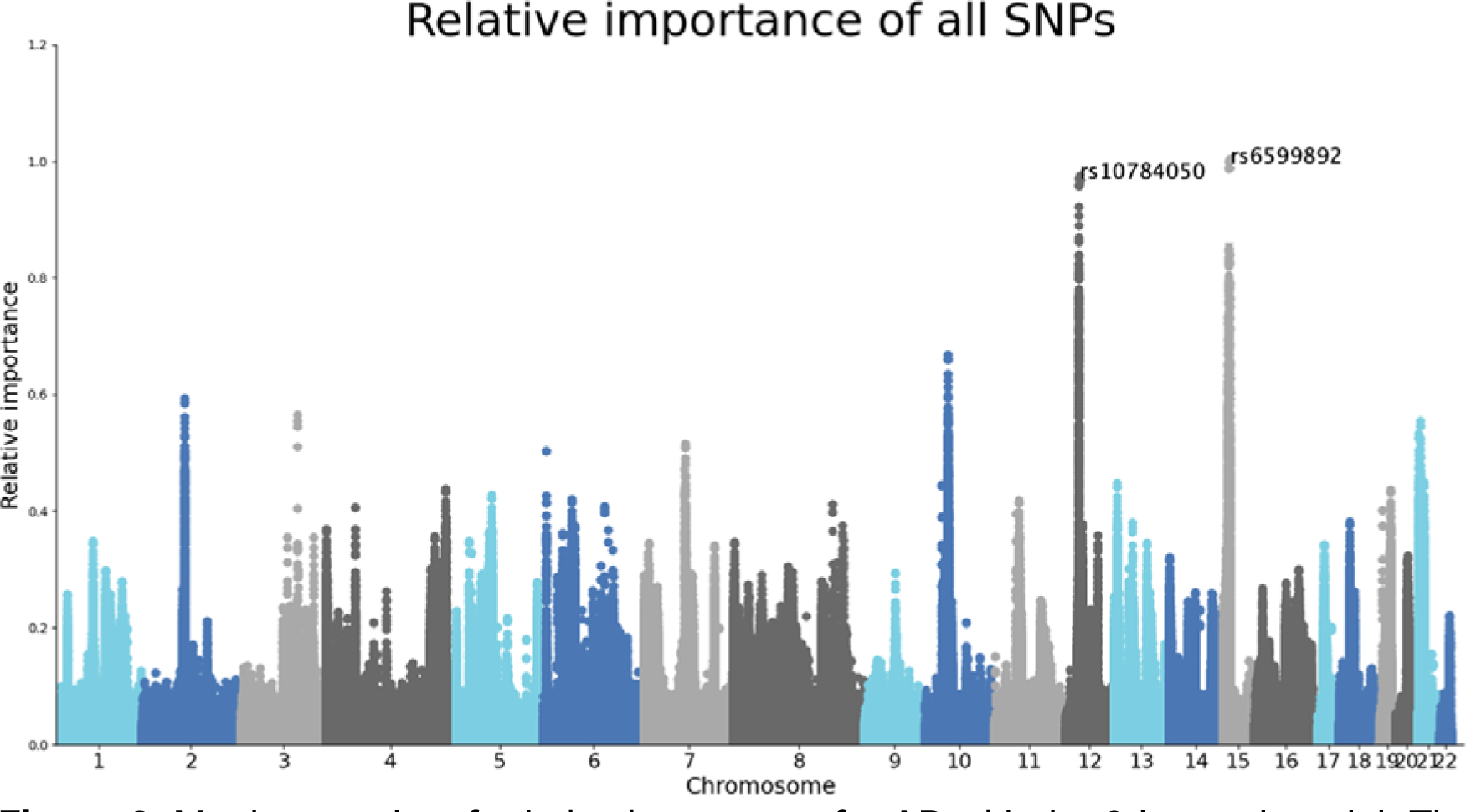
Manhattan plot of relative importance for AD with the 3-layered model. The x-axis shows the chromosomal location of the SNPs and the y-axis the relative importance of the SNPs. The relative importance was calculated for each gene using the learned weights of this neural network.

Based on the sunburst figure, CUB and Sushi multiple domains 1 (*CSMD1)* and RNA binding Fox-1 homolog 1 (*RBFOX1)* genes seem to be significant. *CSMD1* is a gene that is linked with cognitive functions of elderly people. Naturally, it regulates development, connection, and plasticity of circuits of the brain, and is expressed in the central nervous system and epithelial tissues. Several variants in this gene have been reported to be associated with AD, especially in individuals of older age (Stepanov *et al*., 2017). *RBFOX1* has also been previously associated with AD. It is believed to play a role in neuronal development, and in people with AD, it is found around plaques and dystrophic neurites. When the expression of RBFOX1 is lower than normal, the β-amyloid burden increases (Raghavan *et al*., 2020). Also, *RBFOX1* is linked to loss of gray matter in individuals with AD (Vaht *et al*., 2020).C

When using only two layers, the AUC remained relatively high, with 0.73 for the validation set and 0.71 for the training set. The specificity remained high at 0.90 and the sensitivity increased to 0.24.

Ten SNPs were identified from this analysis, at chromosomes 7, 9, 14, 15 and 16. Six of them, rs28488717, rs7186857, rs79654388, rs61711771, rs3901280 and rs4786214, were located at chromosome 16 and were associated with the gene *RBFOX1*, that was identified at the analysis including both layers of genes and pathways. For the rest of the SNPs, the model did not manage to assign a gene. The rs4720976 is associated with PHD finger protein 14 (*PHF14)* (Open Targets Genetics, 2023). There have been no indications that this gene is linked to AD, but it has been hypothesized that age-related diseases of the retina, where *PHF14* is sometimes overexpressed, might be associated with neurodegenerative diseases, such as AD, since they undergo similar changes in the neurons and microglia (Öhman *et al*., 2018). Based on its location to the nearest gene, the family with sequence similarity 74 member A4 (*FAM74A4)* is a potential target gene for rs28670889 (NCBI, 2023). It has been computationally predicted that *FAM74A4* plays a role in neurodegenerative diseases, such as Parkinson’s disease and amyotrophic lateral sclerosis (Haenig *et al*., 2020). Two genes are close to the rs72488234 variant that are of interest (NCBI, 2023). One of them is the olfactory receptor family 11 subfamily H member 12 (*OR11H12)* gene, which is likely associated with differentially hydroxymethylated regions. These are associated with AD pathology, especially through neuritic plaques, which are a histological trait of AD (Zhao *et al*., 2017). The other one is the BCL2 interacting protein 3 pseudogene 6 (*BNIP3P6*) gene (NCBI, 2023). *BNIP3P6* interacts with BCL2, which in turn modulates Ca^+^ signaling in the neurons. The dysregulation of this signaling contributes to the pathogenesis of AD (Callens *et al*., 2021). The rs12594050 variant potentially interacts with *RHPN2P1* based on their distance, a gene that was also identified in the three-layered analysis (NCBI, 2023).

## Discussion

In this study, the performance of traditional GWAS using PLINK was compared to a novel machine learning method, GenNet, which is based on deep neural networks. Genotypic and phenotypic data from the UK Biobank was used to assess the tools. The methods were tested on AD, a binary phenotype. This study was also able to identify potential genes linking new variants to AD.

Through traditional GWAS, four SNPs were found to be significantly associated with AD with the 1 case to 1 control study design and six with the 1 case to 2 controls study design. These variants were located on a chromosomal region that is highly relevant to AD, including genes such as *APOE* and *NECTIN2*. Our analysis also highlighted another gene, *BCL3,* which has also been previously associated with late-onset familial AD and other cognitive impairment-related diseases (Nho *et al*., 2017). The fact that the GWAS analysis using PLINK identified highly plausible genes, indicated that the cohort and epidemiological model used were appropriate and that the comparison with GenNet results would be possible.

The GenNet model trains its neural networks based on prior biological knowledge, reducing the number of trainable parameters (van Hilten *et al*., 2021). This model performed well in the AD binary phenotype, identifying a total of twelve SNPs on seven different biologically plausible genes, while obtaining an AUC of 0.80 when using the SNP, gene annotation and pathway layers, and an AUC of 0.73 when using only the SNP and gene annotation layers.

For the three-layered analysis of the binary trait, GenNet indicated two SNPs associated with two genes which were not previously linked to AD, but with potential biological plausibility. The two genes that were identified, *ALG10B* and *RHPN2P1* are very likely to contribute to AD, as they play important functional roles in cells, such as microglia and neurons (Monasor *et al*., 2020; Parcerisas *et al*., 2014; Schmidt *et al*., 2022). Ten SNPs and five more genes were detected to play a role in AD when using only the SNP and gene annotation layers in the model. Even genes that were not previously identified for AD, such as *BNIP3P6*, are biologically plausible (Callens *et al*., 2021).

As there was no independent set available to run a classification analysis for AD results derived from PLINK to extract an AUC, GenNet results were compared to AUCs reported in the bibliography. AUC from conventional GWAS is reported to be around 75% to 84% (Baker & Escott-Price, 2020). GenNet achieved an AUC of 80% for AD with three layers, and 73% for AD with two layers.

Although the study yielded results, there are several limitations. Firstly, this study model did not take into account the age of AD onset. This is important as AD patients with different onset, for example, early or late, tend to have slightly different genes contributing to their pathology (Hoogmartens, Cacace & Van Broeckhoven, 2021). Moreover, this study did not account for medication. Such modification of the covariates could have increased the explained variance of the models.

An important aspect of GenNet is the freedom for the researcher to assign the network layers. While this introduces ample amount of biological knowledge and flexibility to the models, there is no standard way of doing it, as one may use different algorithms to assign a SNP to a gene and a gene to pathway or use GO terms of different hierarchical levels. It should also be mentioned that several SNPs had no gene linked to them and were left as singletons. When this information is lacking, a gene can be assigned to a SNP based on distance. This method is not very accurate, as SNPs in non-coding regions may interact with genes very far away from where they are located. In fact, they can be up to 2 Mbps apart (Brodie, Azaria & Ofran, 2016).

Another limitation of this study is the way the two approaches were compared. The comparison was mainly based on finding common SNPs and implicated genes between the two methods, and on looking at performance scores of studies of different sample sizes from existing bibliography. However, it would have been more accurate if another comparison method was followed that would, for example, allow us to compare directly AUC scores after also running a classification analysis in PLINK, or use genetic risk scores instead.

To infer associations between the SNPs identified in this study and the phenotype studied, repetition and validation of the analysis is needed. This study should be repeated with, preferably, more data, and taking into account important parameters, such as age of AD onset, that were not considered in the current study. Moreover, the two models should be compared in a more systematic way, such as having an independent set and running classification for AD results derived from PLINK to generate a polygenic risk score and corresponding AUC, and be able to do a direct comparison.

The study identified significant associations between AD and seven different SNPs linked to four different genes from the two study designs using PLINK, and twelve SNPs on seven different genes using GenNet. GenNet obtained an AUC of 0.80 for the three-layered model and of 0.73 for the two-layered model. This is one of the first studies attempting to compare the traditional GWAS approach to machine learning methods using the same data. Overall, this research is one of the few of its kind and more studies are expected in the future performing systematic comparisons to evaluate the advantages and disadvantages of machine learning methods for prediction and understanding of the genetic architecture of complex diseases.

## Data Availability

All data produced in the present study are available upon reasonable request to the authors

## Acknowledgments

This research has been conducted using data from UK Biobank, a major biomedical database (www.ukbiobank.ac.uk). We thank all participants.

